# Unlocking Access: Experience of Discrimination and Differential Care Surrounding Trauma and Acute Care Surgery in People Living with Human Immunodeficiency Virus in a Middle-Income Country

**DOI:** 10.1101/2025.07.18.25331306

**Authors:** Gabriela Zavala Wong, Marianne Luyo, Kirsten Senturia, Larissa Otero, Giuliano Borda Luque, Manuel A. Rodriguez Castro, Eduardo Huamán Egoavil, Stephanie Wilson, Lacey N. LaGrone

## Abstract

**Background:** This observational study aims to identify areas of delayed and differential care, and describe perceived challenges to accessing surgery for people living with human immunodeficiency virus (PLHIV).

**Methods:** An exploratory sequential, observational mixed-methods study, conducted from 2022-2023, in a public hospital in Lima, Peru, included semi-structured interviews with PLHIV and surgeons that informed a survey to describe their perspectives on access to surgical care.

**Results:** Fifteen PLHIV and seven surgeons were interviewed, and 118 surgeons were surveyed. Results were organized by: 1) access to care, 2) patient disclosure, 3) differential treatment for PLHIV, and 4) improvement strategies. PLHIV perceived delayed care resulting from unclear additional presurgical requirements leading to stigma and unexpected costs. Refusal of care made PLHIV feel rejected, disrespected, and discriminated against. For surgeons, mandatory HIV testing was perceived as controversial and beneficial but problematic due to lack of lab resources. PLHIV reported their willingness to disclose was related to their understanding of their disease, particularly knowledge regarding disease transmission. Few surgeons interviewed perceived they provided differential treatment for PLHIV, however, in surveys when comparing surgical practices on PLHIV vs. non-PLHIV (such as double gloving, use of mask/goggles and avoiding extra collaborators while operating), there was a significant difference (p<0.05). Both interviews and surveys revealed that providers educational interventions could potentially reduce differential care and surgery delays.

**Conclusions:** Unclear presurgical requirements delay treatment and lead to unexpected costs. Educational interventions across all specialties, not just surgery, could reduce stigma and help fostering a more inclusive and safer environment for PHLIV.

## INTRODUCTION

Human immunodeficiency virus (HIV) remains a significant global public health problem, accounting for 40.1 million deaths since the virus was first identified in 1981 [1]. HIV has continued to disproportionately affect racial and ethnic minorities, sexual and gender minorities, and low-income groups [1,2], widening the gap in health care access and outcomes [1,3].

Furthermore, people living with HIV (PLHIV) not only face the barriers of a chronic infectious condition but also confront substantial challenges with the overwhelming and growing global burden of surgical problems [4]. For example, in PLHIV cases of low CD4+ count, opportunistic infections have resulted in delayed access to surgery [5,6]. In some instances, HIV positive status also presents a barrier to specific surgical procedures, such as organ donation and reception [6], cardiac surgery [7], bariatric surgery [8,9], and spine surgery [10]. Additionally, within certain communities, the attitudes and actions of healthcare providers have been linked to HIV-related stigma associated with discrimination based on sexual orientation, gender identity and race [11–15]. A systematic review from 2018 evaluated providers’ perspective towards PLHIV, revealing that unsubstantiated fear of acquiring HIV through health care practices reduced quality of care or led to increased frequency of refusal of care. This was especially common among non-HIV specialty providers or providers that had not received HIV stigma training in the past 12 months [16]. Further, a comprehensive multi-site program evaluation suggests that a cycle of poor healthcare-seeking behavior among PLHIV can be perpetuated by PHLIV feeling disliked by, or anticipating shaming from, providers [17].

The lack of literature evaluating the perspectives of PLHIV and providers on access to surgical care is striking. While most studies focus on surgical outcomes, they often overlook the critical issue of equitable access to surgery among this population. In 2016, a study in sub-Saharan Africa showed that organ transplantation is delayed or avoided for PHLIV, highlighting how provider’s concerns about their own increased risk for occupational acquisition of HIV impacts patient receipt of timely surgical care [18].

In response to this existing gap, we conducted an observational mixed-methods study to understand the experiences of PLHIV when accessing acute surgical care, in a limited health system, such as Peru [19].

## MATERIAL AND METHODS

### Setting

Peru, as part of the Andean Region, is an upper-middle-income country with a population of 34 million [20]. According to a global access to surgical modeling study in 2015, an estimated 20.4 million people (60%) in Peru lack access to affordable, safe, and timely surgery [21]. The prevalence of HIV in Peru is 0.4%, affecting 1.4 million people ages 15-49 years old. This rate is lower than the global prevalence of 0.6% and that of Latin America and the Caribbean, which stands at 0.5% [22]. However, Lima (Peru’s capital city) shares the largest HIV burden, accounting for nearly half (44%) of HIV cases in Peru [23]. HIV care in Peru is delivered by health providers from Ministry of Health (MoH) system, the social security health system under the Ministry of Labor system (MoL), the HIV Committee for Uniformed Institutions under the Ministry of Interior, and private sector. Antiretroviral therapy free of cost to patients was available since 2004, initially through the Global Fund, and it is now state funded. Cayetano Heredia Hospital, the research site for interviews in this project, is the MoH institution that delivers most services for PLHIV in Lima, Peru [24].

### Study design and Research Personnel

This observational mixed-methods study, conducted from 2022-2023, included Spanish language interviews and surveys to describe patients’ and providers’ perspectives on PLHIV’s access to surgical care in a public hospital in Lima, Peru. Current literature and the Consolidated Framework for Implementation Research (CFIR) informed [25] the development of semi-structured interview guides (Appendix A and B), the results of which informed survey questions (Appendix C) in an exploratory sequential mixed-methods design. Interviews and surveys included demographic and content questions related to surgical care of PLHIV, possible barriers and facilitators during the providers’ and patients’ encounters, perceived quality of surgical care and overall improvement strategies.

This study was approved by the Institutional Review Boards (IRB) at Universidad Peruana Cayetano Heredia (Project 206718) and at Cayetano Heredia Hospital (Project 059-021). All participants completed a written informed consent prior to participation. The research team reimbursed participants’ transportation and snack expenses as relevant, and compensated interviewees’ time with 30 Peruvian Soles (8 USD).

The research team included a research physician (GZW), a medical anthropologist (KS), three general surgeons (GBL, MRC, EHA), one trauma surgeon (LNL), general physician (ML), physician expert in public health and health sciences (LOV) and medical sociologists (SW). This mixed-methods study took place at a community-based, academic-affiliated public hospital (Cayetano Heredia Hospital) in Lima, Peru.

### Data tools

#### Interviews

Using a purposive sample, semi-structured confidential interviews were conducted over five months. Patients were eligible to participate if they were (1) at least 18 years old, (2) had an HIV diagnosis before surgery, and (3) had undergone or were waiting for a surgical procedure that required admission to operation room (OR). Providers were also recruited for interviews through snowball sampling; they were eligible to participate if they were working as surgeons or residents at Cayetano Hospital surgical service.

Interviews were conducted by a research physician (GZ) in person or via a phone call depending on participants’ availability and, or preference. PLHIV interviews took 20 minutes on average, while surgeons interviews took 9 minutes on average. Informed consent was obtained from all participants prior data collection. Interviews were audio-recorded, transcribed (GZ), translated by a professional medical translator, and audited for accuracy (GZ).

Data sufficiency was determined by GZW, ML and KS and was achieved with 22 interviews [26]. Codebook development was informed by interview transcripts, content expertise and CFIR (multi-step approach was employed), allowing both inductive and deductive codes. Transcripts were coded (GZ, ML, SW) to identify primary themes in Dedoose (version 9.2.012 SocioCultural Research Consultants, LLC), with expert auditing of coding (KS) and following literature standards [27–28]. Thirty percent of interviews were dual coded for 100% agreement that informed the first iteration of codebook development. The thematic analysis served as the basis for survey development. This study conforms to the Standards for Reporting Qualitative Research [29].

#### Surveys

A survey informed by patient and provider interviews, current literature, and CFIR was distributed among surgeons. We applied snowball sampling to recruit participants through targeted efforts at academic conferences, investigators professional networks (GZW, GBL, EHA, MRC and LNL), Peruvian General Surgical Society’s (PGSS) meetings, server list and diverse media channels. Informed consent was provided prior data collection. Anonymous surveys were distributed digitally over 11 months via Microsoft Forms. Quantitative data are presented in frequency tables and chi-square was employed to test for significance of surgical practices reported by providers. STATA (StataCorp. 2023, College Station, TX) was used for data analysis.

### Results overview

We organized the results according to four major themes: 1) access to care, 2) patient disclosure, 3) differential treatment for PLHIV, and 4) improvement strategies. When reporting findings from the interviews, we designated providers or patients as the interviewees; all survey findings were from providers.

## RESULTS

### Demographics

#### Interview participants

Seven surgical providers were interviewed, most (71%) were general surgery attendings, male (86%) and worked at public urban institutions (86%). Of the 15 interviewed PLHIV, 73% were male with a mean age of 39 (SD 11).

All seven interviewed providers reported seeing HIV patients in the emergency room (ER) with acute surgical conditions at least monthly.

#### Survey participants

We collected 118 surveys. More than 90% of respondents were male general surgeons who worked at mostly public urban hospitals (Table 1).

**Table 1.** Surgical Providers Demographics [Surveys].

| Table 1. Surgical Providers Demographics [Surveys] |  | N = 118 | % |
| --- | --- | --- | --- |
| **Gender** |  |  |  |
|  | Male | 97 | 82% |
|  | Female | 21 | 18% |
| **Institution Location** |  |  |  |
|  | Urban | 108 | 92% |
|  | Rural | 5 | 4% |
|  | Suburban | 5 | 4% |
| **Institution Type\*** |  |  |  |
|  | Private | 17 | 14% |
|  | Public | 97 | 80% |
|  | Armed Forces | 4 | 3% |
|  | Academic | 2 | 2% |
|  | Community | 1 | 1% |
| **Surgical Specialty\*** |  |  |  |
|  | General Surgery | 113 | 80% |
|  | Cardiothoracic | 3 | 2% |
|  | Colorectal | 7 | 5% |
|  | Trauma | 12 | 8% |
|  | Oncological Surgery | 1 | 1% |
|  | Pediatric Surgery | 5 | 4% |
|  | Transplant Surgery | 1 | 1% |
| **Years of clinical practice (after completing specialty)** |  |  |  |
|  | In training | 12 | 10% |
|  | 1 to 5 | 40 | 34% |
|  | 6 to 10 | 11 | 9% |
|  | >10 | 55 | 47% |

\*Participants were allowed to pick more than one option

By contrast to interviewees, surgeons surveyed revealed a smaller average of surgeries performed in PLHIV (Table 2).

**Table 2.** Average Monthly Surgeries Performed on People Living with HIV [Surveys] N=118.

|  |  |  |
| --- | --- | --- |
| **<1** | 62 | 53% |
| **1-3** | 35 | 30% |
| **>3** | 15 | 12% |
| **I don't know/blank** | 6 | 5% |

### Access to care

#### Presurgical requirements

First, during interviews PLHIV reported additional presurgical requirements as a major barrier to accessing care. Barriers resulted partly from unclear requirements which seemed unique to PLHIV creating complicated processes, and from additional pre-surgical tests and personal protective equipment (PPE) requirements.

> *I think there are too many protocols that in most cases demand a lot of time and generate additional costs. Because if I am going to have surgery somewhere else, I still need those results, and while the results come out…I would have to get those done on my own and it also generates additional costs. Therefore, either way a patient like me does not gain anything out of this and more than anything else they waste time, waste money, and also have to deal with discrimination. [patient04]*

#### Refusal to provide care

Second, PLHIV stated that access to care was delayed or hindered when providers refused to treat them and referred them to other, more willing, providers. PLHIV mentioned a lack of proper facilities or equipment as reasons providers refused to operate. PLHIV viewed this as wasted valuable time that led to feelings of rejection, disrespect, discrimination, and persecution. Even when referred to a willing provider, PLHIV still had negative feelings about the previous interaction.

#### Mandatory HIV testing

Surveys showed that most surgical providers performed HIV testing when operating on a patient, regardless of whether the surgery was an emergency (72%) or elective (92%) procedure (Fig 1a., Fig. 1b).

**Fig 1a.**
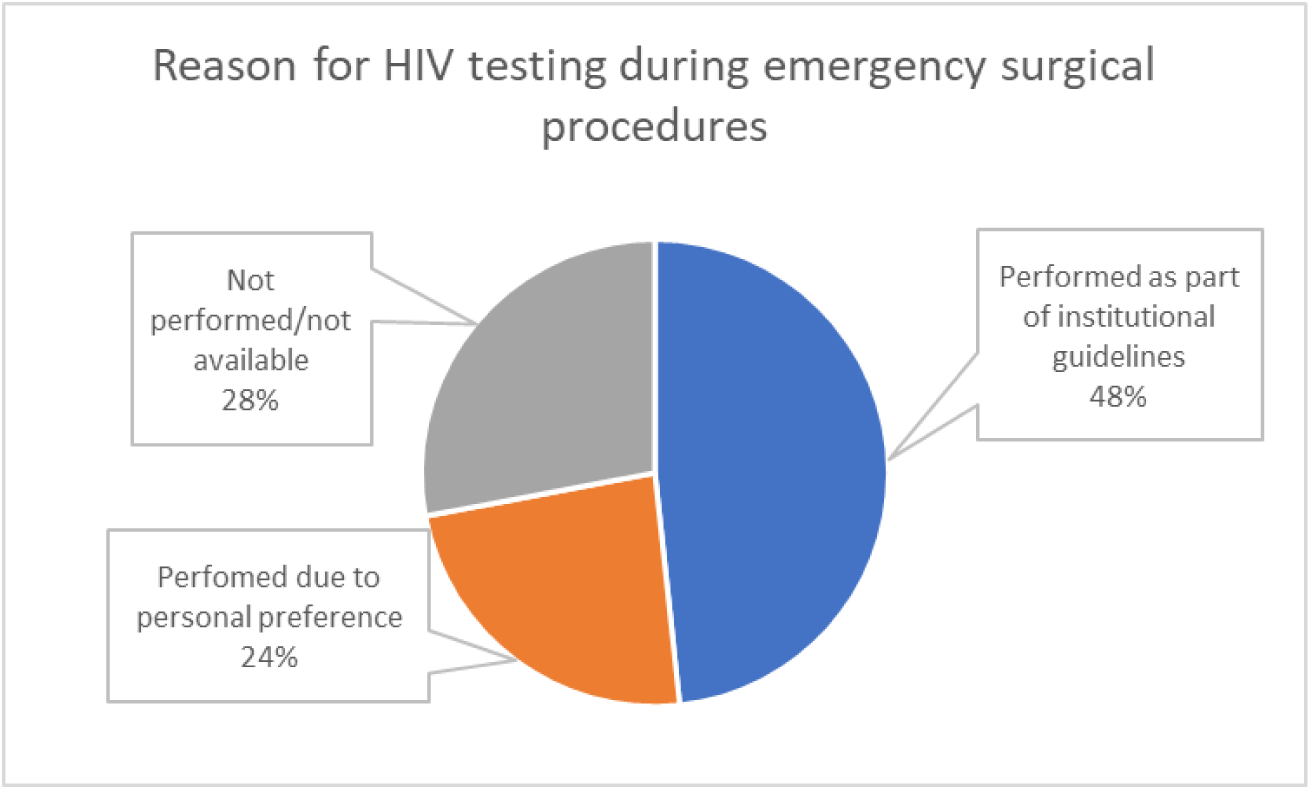
HIV testing during emergency surgical procedures

**Fig 1b.**
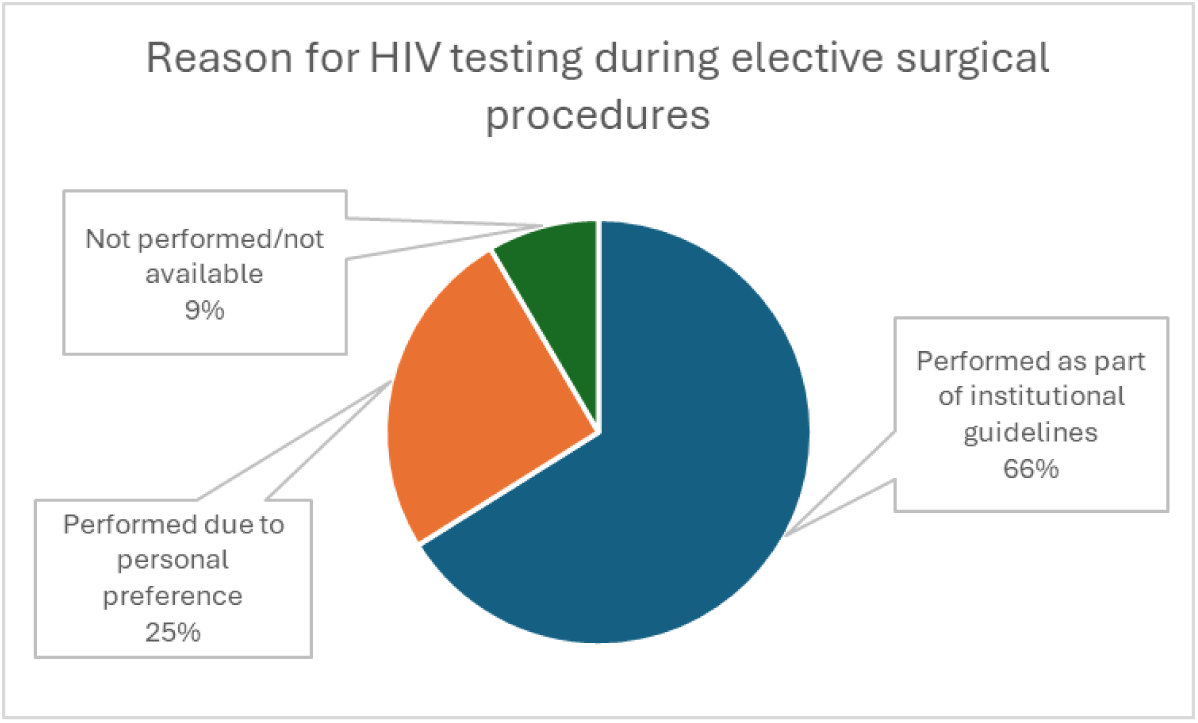
HIV testing during elective surgical procedures

In contradiction to surveyed providers, some interviewed providers declared that mandatory testing was not routinely conducted and was often ignored due to the delays it caused, and the fact that it was dispensable.

> *That would be ideal (to have mandatory HIV testing) in all, in routine patients but unfortunately for issues of economic or the same problems of, that delays and all that, we ignore it. Because the hospital has stock outs of routine laboratory tests. Sometimes there is a lack of lab supplies, I don’t know what. It is a limitation, on that side. [provider01]*

### Patient HIV Disclosure

Interviews with providers regarding PLHIV disclosure of HIV status revealed there was no routine and consistent way that providers discerned patients’ HIV status. However, most providers reported finding out by asking the patient directly.

PLHIV’s willingness to disclose was related to detectability and their perception of their own infectability. For some patients, the willingness to disclose was also related to peace of mind and safety of the doctor.

> *I told him more than everything to avoid discomfort and all that because. I mean, if a person is honest as they say, then they say so, right? Nor does it have to hide because we are all people and I know that they are health professionals and to take more care, but obviously I was already in treatment for almost 6 months and… And I… I knew that I was not a person who could infect or infect. [patient14]*
>
> *I told them that I was an HIV-positive person at all times. I said it for mental health and the peace of mind of the person next to me, especially the doctor. [patient05]*
>
> *But now when I go to this with everything, as a good gesture I do say “just in case I have this”. “Ah good, that you have told us” they tell me. [patient13]*

Positive or “normal” reactions from doctors, whether in the past or anticipated in the present, encouraged patients to disclose HIV status. In the same vein, continued friendliness, patience and “normal” treatment from providers helped PLHIV feel they were receiving the same treatment after HIV disclosure.

PLHIV discussed sometimes feeling obligated to disclose their status. For example, some PLHIV noted providers have the right to know so they can provide the best treatment. Interviews with PLHIV also revealed that the obligation to disclose their HIV status got easier over time after the diagnosis.

Interview results highlighted that while being undetectable gave PLHIV a feeling that they did not need to disclose because they cannot infect others, when providers or intake forms asked, there was still an obligation to disclose.

Other PLHIV mentioned the tendency to omit disclosure because a positive HIV status could interfere with their access to surgery (delays or refusal of care). They also revealed that stigma inhibited disclosure through promoting feelings of embarrassment, fear of rejection, and uncertainty of how, or if, they would be treated by providers.

> *When you mention the pathology, the treatment, the contact is limited, only what you need to know, and if one compares it with other cases, it is different. But as I said before, to prevent those situations I avoid saying that I have this pathology. [patient01]*

In the survey, 88% of providers reported perceiving a patient discomfort when disclosing their HIV status (Table 3).

**Table 3.** Provider Perspective on Patient Discomfort in Disclosing HIV Status [Surveys] N=118.

|  |  |  |
| --- | --- | --- |
| **None** | 5 | 4% |
| **A Few** | 18 | 15% |
| **Some** | 50 | 42% |
| **Most** | 36 | 31% |
| **All** | 7 | 6% |
| **I don't know** | 2 | 2% |

**Table 4.** Providers’ Preferences in The When caring for PLHIV OR [Surveys] (N=118)

| **Table 4. Providers' Preferences in The OR [Surveys] (N=118)** | **When caring for PLHIV** |  | **When caring for non HIV+ patients** |  | **p-value** |
| --- | --- | --- | --- | --- | --- |
| **Moving slowly with more care in the operating room** |  |  |  |  | 0.10 |
|  | 6 | 5% | 11 | 9% |  |
| Rarely | 16 | 14% | 22 | 19% |  |
| Sometimes | 27 | 23% | 25 | 21% |  |
| Usually | 34 | 29% | 24 | 20% |  |
| Always | 35 | 30% | 19 | 16% |  |
| Declined to answer/Blank | 0 | 0% | 17 | 14% |  |
| **Double gloving** |  |  |  |  | <0.05 |
| Never | 8 | 7% | 14 | 12% |  |
| Rarely | 10 | 8% | 27 | 23% |  |
| Sometimes | 21 | 18% | 26 | 22% |  |
| Usually | 26 | 22% | 19 | 16% |  |
| Always | 52 | 44% | 19 | 16% |  |
| Declined to answer/Blank | 1 | 1% | 13 | 11% |  |
| **Non-residents or medical students in the room** |  |  |  |  | 0.06 |
| Never | 9 | 8% | 20 | 17% |  |
| Rarely | 24 | 20% | 21 | 18% |  |
| Sometimes | 36 | 31% | 32 | 27% |  |
| Usually | 21 | 18% | 19 | 16% |  |
| Always | 27 | 23% | 13 | 11% |  |
| Declined to answer/Blank | 1 | 1% | 13 | 11% |  |
| **Use of electric scalpel to replace conventional scalpel** |  |  |  |  | 0.62 |
| Never | 11 | 9% | 14 | 12% |  |
| Rarely | 22 | 19% | 23 | 19% |  |
| Sometimes | 29 | 25% | 25 | 21% |  |
| Usually | 22 | 19% | 22 | 19% |  |
| Always | 34 | 29% | 22 | 19% |  |
| Declined to answer/Blank | 0 | 0% | 12 | 10% |  |
| **Use of mask or goggles** |  |  |  |  | <0.05 |
| **Never** | 2 | 2% | 18 | 15% |  |
| Rarely | 17 | 14% | 28 | 24% |  |
| Sometimes | 28 | 24% | 14 | 12% |  |
| Usually | 21 | 18% | 19 | 16% |  |
| Always | 50 | 42% | 25 | 21% |  |
| Declined to answer/Blank | 0 | 0% | 14 | 12% |  |
| **Avoid extra collaborators in the operating room** |  |  |  |  | <0.05 |
| Never | 4 | 3% | 15 | 13% |  |
| Rarely | 21 | 18% | 39 | 33% |  |
| Sometimes | 36 | 31% | 17 | 14% |  |
| Usually | 25 | 21% | 22 | 19% |  |
| Always | 32 | 27% | 12 | 10% |  |
| Declined to answer/Blank | 0 | 0% | 13 | 11% |  |

### Differential treatment for PLHIV

#### Evidence that treatment is differential

During interviews, providers reported they adopted more PPE (protective glasses and double gloving) in response to knowing a patient in the OR had HIV. When surgeons were surveyed to compare their perceptions on surgical practices when operating on PLHIV vs. non-PLHIV, there was a significant difference (p<0.05) in the reported use of double gloving, use of mask or goggles and avoidance of extra collaborators (assistants) in the operating room.

#### Provider precautions

Similarly, PLHIV stated that they may receive differential treatment in that providers may wear extra PPE; they mentioned some providers may request extra evaluations, such as from the infectious disease service; and that providers described how the existence of extra decontamination requirements in the OR for PLHIV led to delays for surgery.

Providers expressed concerns about possible infection from PLHIV, mentioning possible complications with PLHIV required more concentration and biosecurity measures. These concerns were reinforced when some providers mentioned suffering sharp injuries in the past, leading to behavioral changes such as increased awareness of needles in the OR and changes in surgical equipment to avoid subsequent injuries. Similar results were found in surveyed providers, where 51% stated that they changed the usual OR biosecurity precautions after an exposure to bodily fluids when operating a PLHIV. On a similar note, the survey revealed that only 55% of surgeons report on their institutions providing the PPP they prefer when operating on PLHIV.

Lastly, a subset of providers interviewed mentioned their preference to use a laparoscopic approach to surgery with PLHIV to limit exposure to fluids and secretions. A similar trend was reflected in the survey results, where 59% of providers on a regular basis preferred laparoscopic procedure in PLHIV.

#### Provider reliance on misinformation

Providers also discussed how misinformation among health personnel lead to surgical delays as a form of discrimination against PLHIV. Survey results showed that only 61% (71), highlighted scientific literature as the best source of clinical guidance when operating on PLHIV. Additionally, when surveyed participants were asked about their knowledge of the approximate risk of contracting HIV from exposure of patients’ fluids to their mucosa and from a percutaneous (needle) exposure, just over 30% (38 out of 118) answered 0.09% and 0.33% respectively.

#### Providers determined to avoid differential treatment

While many providers shared examples of differential treatment, others were insistent that PLHIV received the same routine care and treatment as any other patient. Some argued they should treat all patients as if they had HIV, eliminating the possibility of differential treatment. For example, double gloves should be a habit for all patients, so sensitivity was the same for all surgeries performed; in surveys when providers were asked whether double gloving compromised their ability to operate, fewer than 20% reported this as a common concern. During interviews, some providers distinguished between “treatment,” which they reported as consistent across HIV status, and “security,” which they saw as needing to be increased with PLHIV.

### Improvement strategies

#### Training and education

Throughout interviews, PLHIV considered there was a connection between discriminatory care and providers’ lack of adequate and accurate information about HIV as a medical condition. PLHIV saw educational activities as a means of improving their care and reducing discrimination. Some PLHIV were particularly informed and educated about their disease and requirements for care, which in turn empowered them to both anticipate and reject discrimination from providers.

Providers also advocated for education as an improvement strategy. Specifically, providers said that more education about HIV transmission might mitigate the delays for surgery experienced by PLHIV. Survey data confirmed that training and education, specifically about transmission, would be key to improving surgical care access. Out of the 244 answers provided by 118 participants, 68% emphasized the need for education (Figure 2).

**Figure 2.**
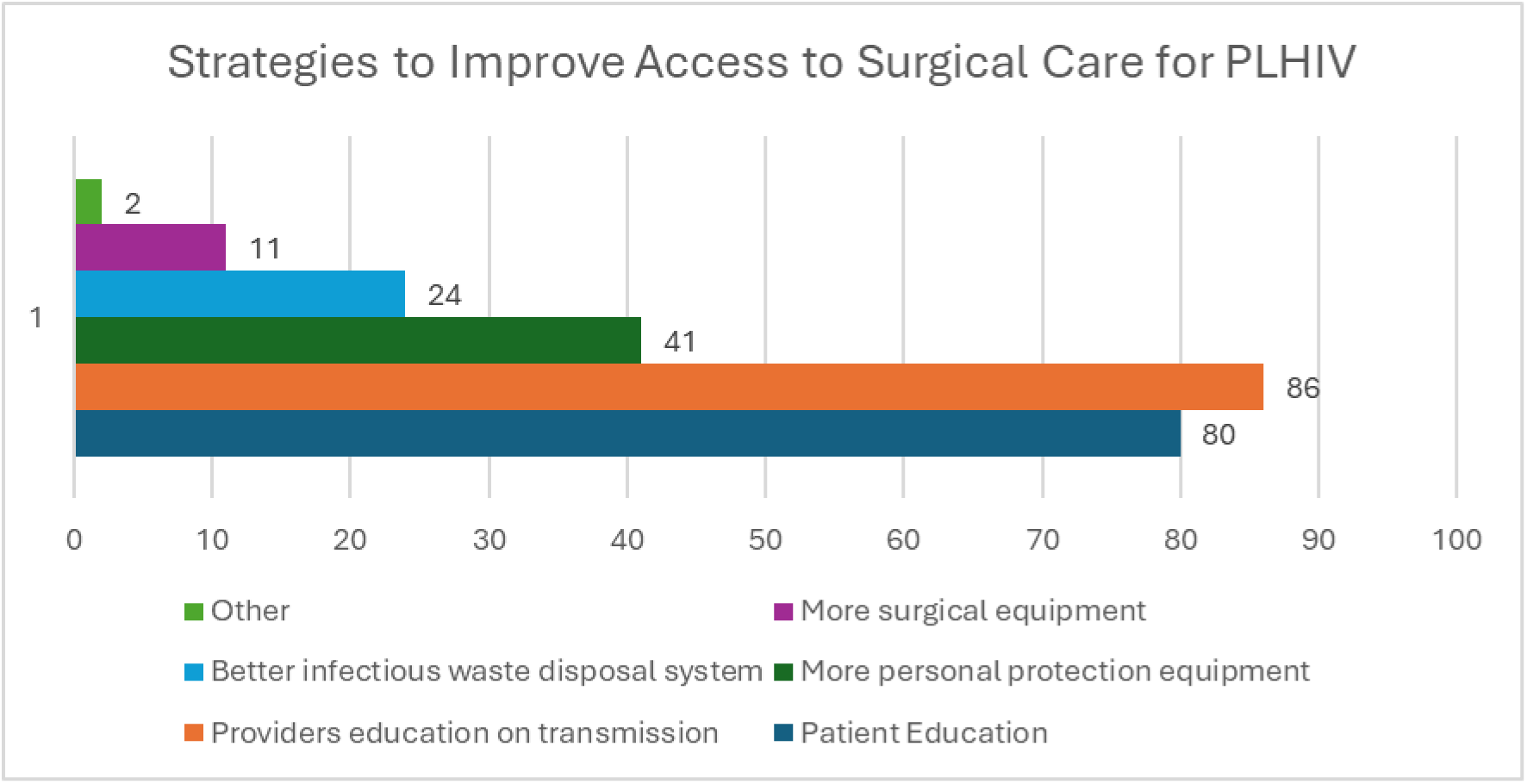
Improvement Strategies

## DISCUSSION

Our mixed methods study found that PLHIV in Peru face barriers in accessing surgical care and perceptions of differential treatment, often feeling uncomfortable in their interactions with surgeons. PLHIV experienced delayed care from additional presurgical requirements, leading to stigma and unforeseen costs. Refusal of care was reported to impede access, causing feelings of rejection, disrespect, and discrimination. Additionally, these influenced patients’ willingness to disclose their HIV status in what felt like an unsafe environment. Nonetheless, it is important to recognize that HIV status disclosure was also affected by the time of HIV diagnosis, as results may suggest that some PLHIV felt more comfortable after some time.

The National Institute for Health Care Excellence (NICE) does not include HIV testing in their routine preoperative tests [30], possibly because of their low HIV prevalence and high viral suppression in the UK [31]. Some Peruvian institutional guidelines do not include it either [32,33]. In 2014, a Peruvian study evaluated the concordance and cost impact of preoperative evaluation tests conducted in a public hospital compared to evidenced-based NICE guidelines and found an inappropriate use of resources (tests that were required even when not supported by evidence) and increased cost of 408,447 USD [34]. Another cost-effectiveness analysis conducted in maxillofacial surgery evaluated the financial implications of HIV screening in patients undergoing elective oral and maxillofacial surgery and suggested that cost per positive HIV resulted higher at 11, 895.3 USD compared to 1,873.8 USD for Hepatitis B and 905.8 USD for Hepatitis C. This suggests that while screening is important, mandatory HIV testing may be tailored to local prevalence [35]. This aligns with 2024 WHO Consolidated Guidelines on Differentiated HIV Testing Services, which suggests that in low HIV burden settings, HIV testing should be offered in clinical settings to individuals with symptoms or medical conditions suspicious of HIV infection, including presumed and confirmed tuberculosis cases [36].

When comparing surgeons reports on surgical practices used with PLHIV vs. non-PLHIV, we found a significant difference (p<0.05) in the reporting of use of double gloving, use of mask or goggles and avoiding extra collaborators in the operating room. However, existing WHO Guidelines for Safe Surgery do not give specific recommendations regarding the HIV-status of the patient, instead they suggest infection prevention and control measures regarding PPE as universal precautions [37]. To reduce stress with occupational risk exposure to HIV infection, providers should be aware that current literature demonstrates the undetectable viral load equals untransmutable (U=U) [38]. This could be the focus of future educational interventions for healthcare providers. However, it is important to recognize that stress associated with the risk of occupational exposure to HIV infection is also affected by the level of viral load suppression and access to post-exposure prophylaxis. Both factors vary depending on the population and the setting and were not addressed in this study.

Finally, educational interventions targeting healthcare providers have proven effective in reducing the HIV epidemic by increasing PLHIV access to care, ensuring timely treatment and overall well-being [39, 40]. Empowering PLHIV can help reduce bias, improve healthcare attitudes and foster greater acceptance [41].

### Limitations

This is an exploratory mixed-methods study that employed non-probability sampling and included interviews with participants in a single urban setting, which limits our ability to generalize to more rural or suburban environments. Furthermore, the interview guide focused primarily on patient’s access to surgical care, and we recognize there are other areas of healthcare where PLHIV may encounter and suffer stigma and discrimination that were outside the scope of this project. People not living with HIV were not interviewed regarding their perceptions on stigma, discrimination, or difficulties when accessing care. Many individuals, including those without HIV, already encounter unmet medical needs, long waiting times, and a lack of infrastructure based on the 2025 Organisation for Economic Co- operation and Development (OECD) review on Peruvian health system [19]. Our results could shed light on deficiencies experienced not only by PLHIV, but it is important to acknowledge that the historical stigma associated with HIV may further widen the gap in accessing care.

## CONCLUSION

Our study shows that stigma against PLHIV remains a significant issue in surgical care environments in hospitals in Lima. Unnecessary testing and unclear presurgical protocols not only delay treatment but may also lead to inefficient use of resources. Identifying the challenges for both patients and providers may contribute to the development of transformative interventions and policies to empower patients to seek the care they need. Furthermore, this study could contribute to developing and implementing targeted training interventions aimed at dismantling HIV-related stigma to equip healthcare surgical providers with the knowledge and skills needed to create a safer environment for patients and surgeons and offer compassionate informed care to PLHIV.

## Data Availability

All data produced in the present work are contained in the manuscript. Supplemental material (as interview guide and survey) is available upon reasonable request to the authors

## ACKNOWLEDGEMENTS

This research project was supported by the National Institutes of Health (NIH) Fogarty International Center of the National Institutes of Health and the Program for Advanced Research Capacities for AIDS in Peru (PARACAS) under grant #D43TW009345 awarded to the Northern Pacific Global Health Fellows Program. Special thanks to the Sociedad de Cirujanos del Perú and their 2023 executive board for supporting the survey distribution among their members.

## Funding Statement

This research project was supported by the National Institutes of Health (NIH) Fogarty International Center of the National Institutes of Health and the Program for Advanced Research Capacities for AIDS in Peru (PARACAS) under grant #D43TW009345 awarded to the Northern Pacific Global Health Fellows Program.

There are no conflicts of interest to declare by all authors.

Informed consent was obtained from all individual participants included in the study.

Study was approved by Universidad Peruana Cayetano Heredia IRB (206718) and Cayetano Heredia Hospital IRB (059-021)

## REFERENCES

1. https://www.who.int/news-room/fact-sheets/detail/hiv-aids

2. Lightfoot M, Milburn N, Loeb Stanga L. Addressing Health Disparities in HIV: Introduction to the Special Issue. J Acquir Immune Defic Syndr. 2021 Dec 15;88(S1):S1–S5. doi: 10.1097/QAI.0000000000002804. PMID: 34757986; PMCID: PMC8579984.

3. Alexandra Marshall S, Brewington KM, Kathryn Allison M, Haynes TF, Zaller ND. Measuring HIV-related stigma among healthcare providers: a systematic review. AIDS Care - Psychological and Socio-Medical Aspects of AIDS/HIV. 2017;29(11):1337–45.

4. Debas HT, Donkor P, Gawande A, Jamison DT, Kruk ME, Mock CN, editors. Essential Surgery: Disease Control Priorities, Third Edition (Volume 1) [Internet]. Washington (DC): The International Bank for Reconstruction and Development / The World Bank; 2015 [cited 2022 Mar 24]

5. Bowa K, Kawimbe B, Mugala D, Musowoya D, Makupe A, Njobvu M, et al. Send Orders for Reprints to The Open AIDS Journal A Review of HIV and Surgery in Africa. The Open AIDS Journal [Internet]. 2016;10:16–23. Available from: www.benthamopen.com/TOAIDJ/

6. Roland ME, Barin B, Huprikar S, Murphy B, Hanto DW, Blumberg E, et al. Survival in HIV-positive transplant recipients compared with transplant candidates and with HIV-negative controls. Aids. 2016;30(3):435–44.

7. Dominici C, Chello M. Impact of human immunodeficiency virus (HIV) infection in patients undergoing cardiac surgery: a systematic review. Reviews in Cardiovascular Medicine. 2020 Sep 30;21(3):411–8.

8. Pourcher G, Peytavin G, Schneider L, Gallien S, Force G, Pourcher V. Bariatric surgery in HIV patients: experience of an Obesity Reference Center in France. Surg Obes Relat Dis. 2017 Dec;13(12):1990–6.

9. Fysekidis M, Cohen R, Bekheit M, Chebib J, Boussairi A, Bihan H, et al. Sleeve gastrectomy is a safe and efficient procedure in HIV patients with morbid obesity: a case series with results in weight loss, comorbidity evolution, CD4 count, and viral load. Obes Surg. 2015 Feb;25(2):229–33.

10. Farias FAC, Dagostini CM, Falavigna A. HIV and Surgery for Degenerative Spine Disease: A Systematic Review. J Neurol Surg A Cent Eur Neurosurg. 2021 Sep;82(5):468–74.

11. Belenko S, Dembo R, Copenhaver M, et al. HIV stigma in prisons and jails: Results from a staff survey. AIDS Behav 2016;20:71–84. [PMC free article] [PubMed] [Google Scholar]

12. Davtyan M, Olshansky EF, Brown B, Lakon C. A grounded theory study of HIV-related stigma in U.S.-based healthcare settings. J Assoc Nurses AIDS Care 2017;28:907–922. [PubMed] [Google Scholar]

13. Fredericksen R, Edwards T, Crane HM, et al. Patient and provider priorities of self-reported domains in HIV clinical care. AIDS Care 2015;27:1255–1264. [PMC free article] [PubMed] [Google Scholar]

14. Stringer KL, Turan B, McCormick L, et al. HIV-related stigma among healthcare providers in the Deep South. AIDS Behav 2016;20:115–125. [PMC free article] [PubMed] [Google Scholar]

15. Varas-Diaz N, Neilands T, Rivera S, Betancourt E. Religion and HIV/AIDS stigma, Implications for health professionals in Puerto Rico. Glob Public Health 2010;5:295–312. [PMC free article] [PubMed] [Google Scholar]

16. Geter A, Herron AR, Sutton MY. HIV-Related Stigma by Healthcare Providers in the United States: A Systematic Review. AIDS Patient Care STDS. 2018 Oct;32(10):418–424. doi: 10.1089/apc.2018.0114. PMID: 30277814; PMCID: PMC6410696.

17. Magnus M, Herwehe J, Murtaza-Rossini M, Reine P, Cuffie D, Gruber D, et al. Linking and Retaining HIV Patients in Care: The Importance of Provider Attitudes and Behaviors. AIDS Patient Care STDS. 2013 May;27(5):297–303.

18. Bowa K, Kawimbe B, Mugala D, Musowoya D, Makupe A, Njobvu M, Simutowe C. A Review of HIV and Surgery in Africa. Open AIDS J. 2016 Apr 8;10:16-23. Doi: 10.2174/1874613601610010016. PMID: 27347268; PMCID: PMC4893540.

19. Organisation for Economic Co-operation and Development. OECD Reviews of Health Systems: Peru 2025. Paris: OECD Publishing; 2025. 10.1787/f3ddb6a4-en.

20. World Bank. Peru Overview [Internet]. 2024 [cited 2024 Oct 6]. Available from: https://www.worldbank.org/en/country/peru/overview

21. Alkire BC, Raykar NP, Shrime MG, Weiser TG, Bickler SW, Rose JA, Nutt CT, Greenberg SL, Kotagal M, Riesel JN, Esquivel M, Uribe-Leitz T, Molina G, Roy N, Meara JG, Farmer PE. Global access to surgical care: a modelling study. Lancet Glob Health. 2015 Jun;3(6) doi: 10.1016/S2214-109X(15)70115-4. Epub 2015 Apr 27. PMID: 25926087; PMCID: PMC4820251

22. World Bank. Prevalence of HIV, total (% of population ages 15-49) [Internet]. 2024 [cited 2024 Oct 6]. Available from: https://data.worldbank.org/indicator/SH.DYN.AIDS.ZS?locations=SZ

23. Reyes N, Benites C, García-Fernández L, Calderon M, Fiestas F, Vasquez-Becerra R, et al. HIV treatment cascade in regions of Peru with the highest HIV prevalence. HIV Med. 2023 May;24(5):620–7. doi:10.1111/hiv.13452.

24. Ministerio de Salud. Hospital Cayetano Heredia brinda mayor cantidad de tratamientos a personas con VIH/SIDA [Internet]. 2023 [cited 2024 Oct 6]. Available from: https://www.gob.pe/institucion/minsa/noticias/42259-hospital-cayetano-heredia-brinda-mayor-cantidad-de-tratamientos-a-personas-con-vih-sida

25. CFIR Research Team. CFIR Constructs [Internet]. 2024 [cited 2024 Oct 6]. Available from: https://cfirguide.org/constructs/

26. Vasileiou K, Barnett J, Thorpe S, Young T. Characterising and justifying sample size sufficiency in interview-based studies: systematic analysis of qualitative health research over a 15-year period. BMC Med Res Methodol. 2018;18(1):148. doi:10.1186/s12874-018-0594-7

27. Hennink MM, Hutter I, Bailey A. Qualitative research methods. 2nd ed. London: SAGE Publications Ltd; 2020.

28. Pope C, Ziebland S, Mays N. Qualitative research in health care: Analysing qualitative data. BMJ. 2000;320(7227):114–6. 10.1136/bmj.320.7227.114

29. O’Brien, B. C., Harris, I. B., Beckman, T. J., Reed, D. A., & Cook, D. A. (2014). Standards for reporting qualitative research: a synthesis of recommendations. Acad Med, 89(9), 1245–1251. 10.1097/ACM.0000000000000388

30. National Institute for Health and Care Excellence. Routine preoperative tests for elective surgery [Internet]. 2016 Apr 5 [cited 2024 Oct 6]. Available from: https://www.nice.org.uk/guidance/ng45

31. Terrence Higgins Trust. HIV statistics [Internet]. London: Terrence Higgins Trust; [cited 2025 Jul 18]. Available from: https://www.tht.org.uk/hiv/about-hiv/hiv-statistics.

32. Ministerio de Salud del PerU. Resolucion Directoral N° 193-2024-HNCH-DG. Lima: Hospital Nacional Cayetano Heredia; 2024.

33. Instituto Nacional de Salud del Niño San Borja. Resolución Directoral N° 000037-2021-DG- INSNSB: Directiva Sanitaria de Cirugía Ambulatoria. Lima: INSNSB; 2021

34. Buho. Concordancia e impacto en costos entre la evaluación preoperatoria realizada en un hospital de ESSALUD y la guía clínica basada en evidencia de utilización de pruebas preoperatorias para cirugía electiva. Chiclayo: 2014 Sep 26].

35. Sukegawa S, Sukegawa Y, Hasegawa K, Ono S, Nakamura T, Fujimura A, Fujisawa A, Nakano K, Takabatake K, Kawai H, Mukainaka Y, Nagatsuka H, Furuki Y. The effectiveness of pre-operative screening tests in determining viral infections in patients undergoing oral and maxillofacial surgery. Healthcare (Basel). 2022 Jul 20;10(7):1348. doi: 10.3390/healthcare10071348. PMID: 35885875; PMCID: PMC9324129.

36. 3.World Health Organization. Consolidated guidelines on differentiated HIV testing services [Internet]. Who.int. World Health Organization; 2024. Available from: https://www.who.int/publications/i/item/9789240096394

37. World Health Organization. WHO Guidelines for Safe Surgery 2009 Safe Surgery Saves Lives [Internet]. 2009. Available from: https://apps.who.int/iris/bitstream/handle/10665/44185/9789241598552_eng.pdf

38. Centers for Disease Control and Prevention. Undetectable = Untransmittable. Available from: https://www.cdc.gov/global-hiv-tb/php/our-approach/undetectable-untransmittable.html. Accessed 2024 Oct 5.

39. UNAIDS. Eliminating discrimination: guidance. Available from: https://www.unaids.org/en/resources/documents/2022/eliminating-discrimination-guidance_en. Accessed 2024 Oct 5.

40. Nyblade L, Srinivasan K, Mazur A, Raj T, Patil DS, Devadass D, et al. HIV stigma reduction for health facility staff: development of a blended-learning intervention. Front Public Health. 2018;6:165. doi:10.3389/fpubh.2018.00165. Available from: 10.3389/fpubh.2018.00165. Accessed 2024 Oct 5.

41. Ocloo J, Matthews R. From tokenism to empowerment: progressing patient and public involvement in healthcare improvement. [Narrative review]. Available from: [insert journal name and publication details if available]. Accessed 2024 Oct 5.

